# Linking EMS and In-Hospital Stroke Records: Impact of Deterministic vs. Probabilistic Methods on Selection Bias

**DOI:** 10.1101/2025.08.01.25332630

**Authors:** Christopher J McLouth, Larry B Goldstein

## Abstract

**Background:** Linking emergency medical services (EMS) and hospital stroke registry data is crucial for evaluating stroke systems of care, but the impact of different linkage methods on selection bias remains unclear. This study compared deterministic and probabilistic linkage approaches and assessed their effects on sample representativeness and analytical conclusions

**Methods:** In this cross-sectional study we analyzed 13,567 stroke patients transported by EMS to 40 Kentucky hospitals participating in Get With The Guidelines – Stroke (2021-2023). Records were linked using deterministic and probabilistic methods. We compared match rates, assessed sample representativeness, and evaluated the impact of selection bias using inverse probability weighting.

**Results:** Deterministic and probabilistic methods achieved match rates of 73.0% and 78.7%, respectively. Both methods produced similar representative samples, with modest differences between matched and unmatched cases primarily in race and admission year. Accounting for selection bias had minimal impact on the estimated associations between EMS stroke recognition and outcomes (percent change in adjusted odds ratios < 1%).

**Conclusions:** While probabilistic linkage yielded modestly higher match rates, both methods produced comparable results with minimal selection bias. When working with high-quality data with low missingness, deterministic linkage may be sufficient for many analyses, though sensitivity analyses remain important for assessing potential bias.

## Introduction

Timely treatment of patients having an acute ischemic stroke is critical, as delays between symptom onset, hospital arrival, and treatment are associated with worse outcomes. ^1–7^ Prehospital management is an essential component of the overall stroke care system. The American Heart Association’s (AHA) Get With the Guidelines-Stroke® (GWTG-S) program includes quality of care metrics for both pre-hospital and in-hospital stroke patient care. ^8,9^ Other hospital-based quality improvement programs, such as the Paul Coverdell National Acute Stroke Program (PCNASP), have also been established.^10^ Participation in PCNASP is associated with increased adherence to stroke care performance measures, and within this program, higher quality care is associated with lower odds of death and higher odds of discharge to home.^11–13^ There are, however, gaps connecting prehospital care improvement programs with hospital-based metrics and longer-term patient outcomes.^14^ Many states maintain EMS registries based on standardized elements from the National Emergency Medical Services Information System (NEMSIS), which could be leveraged to help guide EMS quality improvement activities.^15^ Connecting the data collected in these registries with hospital-level care databases is challenging because direct linkage of records based on unique identifiers (e.g., name, social security number) is generally not possible due to administrative and legal restrictions. This lack of integrated data systems and absence of unique patient identifiers hinders a comprehensive analysis based on both pre-hospital and hospital activities and their impact on post-stroke outcomes.

Despite the lack of unique identifiers, records across EMS and hospital data sources can be linked using several approaches. Deterministic and probabilistic linkage are two commonly used linkage methods. Deterministic methods require exact agreement on multiple variables such as age, date of birth, sex, and arrival time. This can be done in a single step or through a hierarchical process that applies progressively less restrictive match criteria. In contrast, probabilistic methods use statistical models to estimate the similarity between two records. A popular approach is based on a theoretical framework that uses an expectation-maximization algorithm to generate a linkage score for pairs of records.^16,17^ Compared to deterministic methods, probabilistic linkage can better accommodate missing or inconsistent data, but may yield more false positives and can require manual review.

Both deterministic and probabilistic methods have been used in stroke research and care improvement programs, with matching typically based on age, date of birth, sex, hospital arrival date, and receiving hospital. One study matched 62.8% of stroke patients to an EMS transport registry using deterministic linkage.^18^ Another compared deterministic and probabilistic methods with 45.5% and 62.8% match rates, respectively, with probabilistically linked samples being more representative of the full patient population.^19^

The extent to which the choice of linkage methods impacts downstream analyses and inference remains uncertain. In particular, selection bias introduced by linking only a subset of records may affect the validity and generalizability of results.^20^ Using state-wide data, we aimed to: 1, compare deterministic and probabilistic linkage methods for EMS and in-hospital stroke data; 2, evaluate the representativeness of the linked samples by comparing characteristics and outcomes between matched and unmatched records; and 3, assess how accounting for selection bias influences estimates of the association between EMS care variables and post-stroke outcomes.

## Methods

### In-Hospital and EMS Registry Data

In-hospital data were obtained from Kentucky hospitals participating in the GWTG-S registry, a national, prospective, observational quality improvement initiative sponsored by the AHA and the American Stroke Association (ASA). Participating hospitals collect data on patients hospitalized with acute ischemic stroke, hemorrhagic stroke, or transient ischemic attack using a standardized set of variables. Trained hospital staff abstract clinical and demographic information, treatments, and in-hospital outcomes using a web-based patient management tool. The registry includes detailed information on patient characteristics, medical history, diagnostic testing, time metrics (e.g., door-to-computed tomography (CT) time), treatments (e.g., IV thrombolysis, mechanical thrombectomy), and discharge outcomes. This study includes data from 40 of 97 (41.2%) acute care facilities in Kentucky who participate in GWTG-S. Only de-identified data were used in this analysis.

EMS data were obtained from the Kentucky Board of Emergency Medical Services (KBEMS), which maintains a statewide repository of EMS transportation records submitted by licensed EMS agencies. Data are collected using standardized formats aligned with the National EMS Information System (NEMSIS) and include dispatch details, response times, patient assessments (e.g., stroke scale administration, primary diagnostic impression), physical measures, and transport information. All EMS transports between the scene and the hospital are included in data extractions. This study includes data from 217 EMS agencies in the state of Kentucky.

The time period for the current study was the years 2021 – 2023. The study was approved by the University of Kentucky’s Institutional Review Board.

### Deterministic and Probabilistic Matching Method

Prior to implementing data matching methods, the data were reviewed for consistency and recoded to ensure compatibility between the two data sources. EMS transports with incomplete destination hospital information were recoded to their most likely GWTG-S hospital when possible (e.g., slight misspelling) or left missing. GWTG-S registry data was restricted to patient arrival “EMS from home/scene.”

A deterministic match was performed using sex, date of birth, date of arrival/transfer, and destination location. Records were linked if they were exact matches on all four criteria. To allow for some discrepancy in arrival time, records matching the prior criteria were assumed to be a valid match if the EMS transfer of care time was within 60 minutes of the arrival time documented in GWTG-S. We used a many-to-many matching approach that allowed each GWTG-S record to be matched to multiple EMS transportation records, and vice versa. In cases with multiple potential matches, we retained the pair with the smallest discrepancy in arrival time, which prevented any GWTG-S or EMS record from appearing more than once in the final dataset.

Link Plus software (Version 3.0, Centers for Disease Control and Prevention) was used to perform probabilistic matching by calculating a linkage score based on the same variables as the deterministic match. Fields with exact matches increase the overall score, whereas the score is decreased by mismatches or missing values. Higher scores indicate a greater likelihood of a true match. Candidate matches were evaluated based on the extent and type of discrepancy across the fields. Exact matches were defined as record pairs that agreed on all linkage variables. These met the same criteria used in the deterministic match, and a similar 60-minute arrival time window was used. Inexact matches exhibited minor discrepancies, such as missing values in the EMS record (e.g., sex or date of birth) or slight mismatches in arrival date (e.g., 11/5/2021 vs 5/11/2021) or location (e.g., correct hospital system was recorded but specific branch may be misclassified). For inexact matches, a stricter 20-minute time window was applied to reduce false positives. In cases in which multiple EMS transports matched a single GWTG-S record, the record with the shortest discrepancy in arrival time was retained.

### Data Analysis

For each method, the matching percentage was calculated as the number of EMS records linked to a GWTG-S record divided by the total number of GWTG-S records that were EMS transports. Deterministic and probabilistic percentages were compared via a chi-square test. Next, we assessed the representativeness of each sample for deterministic and probabilistic matches by comparing demographic and clinical characteristics of the matched and unmatched individuals. These variables included age, sex, race, year, stroke type, admission National Institute of Health Stroke Scale (NIHSS), EMS pre-hospital notification, door-to-CT time, and discharge disposition as recorded in GWTG-S. Due to the large sample size, measures of effect were included with p-values when comparing matched and unmatched patients. For continuous variables, a standardized mean difference was calculated on raw values if normally distributed or ranks if non-normally distributed. Standardized differences for categorical variables were calculated using a multivariate Mahalanobis distance.^21^

To estimate potential selection bias arising from incomplete linkage, we applied an inverse probability of selection weighting approach, similar to using propensity score methods to adjust for confounding. Specifically, we modeled the probability of successful linkage using multivariable logistic regression, incorporating patient-level variables collected in GWTG-S that were potentially associated with both linkage success and clinical outcomes. The predicted probabilities from this model represented each patient’s likelihood of being successfully linked. Each linked record was then weighted by the inverse of its predicted linkage probability, creating a pseudo-population in which the distribution of observed clinical and demographic characteristics was similar to that of the full GWTG-S population who were transported by EMS.

We conducted all analyses using both unweighted models, which include only successfully linked cases without adjustment for linkage bias, and inverse probability of selection weighting-adjusted models, which accounted for the probability of linkage. Unweighted and weighted adjusted odds ratios (aOR) were compared within linkage method by calculating the percent change as follows:

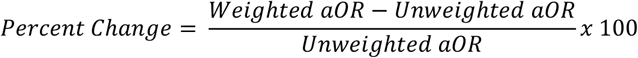

This allowed us to assess the extent to which incomplete linkage may have biased estimates of association. All models included covariates.

To assess the potential impact of selection bias resulting from incomplete linkage, we determined the relationship between EMS stroke recognition and both door-to-CT times and discharge disposition (home vs other). EMS stroke identification was defined as either a primary or secondary impression of the following ICD-10 codes: Stroke (I63.9), Cerebral infarction, stroke (I63), TIA (G45.9), and Cerebrovascular disease, unspecified (I67.9). Door-to-CT time was dichotomized as being less than or equal to 25 minutes or greater than 25 minutes, a quality metric. Discharge disposition was dichotomized as home vs other. These models were estimated using multivariable logistic regression.

SAS version 9.4 (SAS Institute INC, Cary, NC) was used for all data management and statistical analysis. Significance was determined in reference to both the p-value and the standardized difference. Missing data was handled either using listwise deletion or by including a category for missing. This work represents the author’s independent analysis of local or multicenter data gathered using the AHA Get With The Guidelines IQVIA Registry Platform™ but is not an analysis of the national GWTG dataset and does not represent findings from the AHA GWTG National Program.

## Results

### Comparison of Deterministic and Probabilistic Linkage Percentages

Between January 1, 2021 and December 31, 2023, 13,567 stroke patients arrived via EMS to one of the 40 Kentucky hospitals submitting data to GWTG-S. For the deterministic match, 9,898 EMS records were an exact match on sex, date of birth, date of arrival/transfer, and destination location with documented arrival times different by less than 60 minutes, resulting in a match rate of 73.0%. The probabilistic match method identified an additional 780 patients (n = 10,679), resulting in a match rate of 78.7%. This increase of 5.7% was significant (χ^2^(1) = 122.67, p <.001).

### Representativeness of Matched Samples

Table 1 compares the demographic and clinical characteristics of successfully matched stroke cases to unmatched cases within each matching strategy. For the deterministic matching strategy, race and year of presentation had the most notable differences. A higher proportion of unmatched patients were White (89.9% vs. 85.1%; standardized difference = 0.147; p < 0.001). Admission year also differed (standardized difference = 0.062; p < 0.001), with match rates decreasing from 2021 to 2023. There were differences in admission NIHSS scores (median 6 vs. 5; standardized difference = 0.067; p = 0.001) and NIHSS category distribution (p = 0.004; standardized difference = 0.032), although only the continuous NIHSS score reached a potentially meaningful standardized difference. Other variables, including age, sex, insurance status, stroke type, and key treatment variables such as IV thrombolysis and door-to-CT time, showed no differences (standardized differences < 0.05), indicating the matched sample was representative of the full sample on these variables.

**Table 1.** Comparison of Matc hed and Unmatched Samp les Under Determ inistic and Pr obabili stic Match ing Approaches.

|  | Deterministic match |  |  |  | Probabilistic Match |  |  |  |
| --- | --- | --- | --- | --- | --- | --- | --- | --- |
|  | Match<br>(n = 9,898) | Unmatched<br>(n = 3,669) | Standardized<br>Difference | p-value | Matched<br>(n = 10,679) | Unmatched<br>(n = 2,888) | Standardized<br>Difference | p-value |
| **Age, mean (SD)** | 70.8 (14.0) | 70.3 (14.0) | 0.035 | 0.067 | 70.7 (14.0) | 70.4 (13.9) | 0.022 | 0.292 |
| **Sex, n (%)** |  |  |  |  |  |  |  |  |
| Female | 5194 (52.5%) | 4704 (51.8%) | 0.015 | 0.195 | 5594 (52.4%) | 1497 (51.9%) | 0.011 | 0.765 |
| **Race, n (%)** |  |  |  |  |  |  |  |  |
| White | 8419 (85.1%) | 3299 (89.9%) | 0.147 | <.001 | 9102 (85.2%) | 2616 (90.6%) | 0.164 | <.001 |
| **Insurance Status** |  |  | 0.012 | 0.481 |  |  |  |  |
| Medicare | 6240 (70.5%) | 2341 (70.8%) |  |  |  |  |  |  |
| Non-Medicare | 2608 (29.5%) | 964 (29.2%) |  |  |  |  |  |  |
| Missing | 1050 (10.6%) | 364 (9.9%) |  |  |  |  |  |  |
| **Stroke type** |  |  | 0.01 | 0.721 |  |  | 0.025 | 0.253 |
| Ischemic<br>Stroke/TIA | 3148 (85.9%) | 8448 (85.4%) |  |  | 9106 (85.3%) | 2490 (86.3%) |  |  |
| Hemorrhagic<br>Stroke | 495 (13.5%) | 1389 (14.0%) |  |  | 1509 (14.1%) | 375 (13.0%) |  |  |
| Stroke NOS/No<br>Stroke Diagnosis | 57 (0.6%) | 22 (0.6%) |  |  | 60 (0.6%) | 19 (0.7%) |  |  |
| **Year** |  |  | 0.062 | <.001 |  |  | 0.072 | <.001 |
| 2021 | 3296 (33.3%) | 1056 (28.8%) |  |  | 3533 (33.1%) | 819 (28.5%) |  |  |
| 2022 | 3367 (34.0%) | 1265 (34.5%) |  |  | 3661 (34.3%) | 971 (33.8%) |  |  |
| 2023 | 3235 (32.7%) | 1335 (36.4%) |  |  | 3485 (32.6%) | 1085 (37.7%) |  |  |
| **Admission**<br>**NIHSS Score,**<br>**median [IQR]** | 5 [2 - 13] | 6 [2 - 14] | 0.067 | 0.001 | 5 [2 - 13] | 6 [2 - 14] | 0.054 | 0.014 |
| **NIHSS Category** |  |  | 0.032 | 0.004 |  |  | 0.034 | 0.004 |
| 0-5 | 4663 (47.1%) | 1599 (43.6%) |  |  | 5000 (46.8%) | 1262 (43.7%) |  |  |
| 6-10 | 1740 (17.6%) | 648 (17.7%) |  |  | 1875 (17.6%) | 513 (17.8%) |  |  |
| 11-15 | 984 (9.9%) | 403 (11.0%) |  |  | 1047 (9.8%) | 340 (11.8%) |  |  |
| 16-20 | 707 (7.1%) | 283 (7.7%) |  |  | 760 (7.1%) | 230 (8.0%) |  |  |
| >20 | 1149 (11.6%) | 450 (12.3%) |  |  | 1270 (6.8%) | 329 (11.4%) |  |  |
| Missing | 655 (6.6%) | 286 (7.8%) |  |  | 727 (6.8%) | 214 (7.4%) |  |  |
| **IV Thrombolytic, n (%) Yes** | 1296 (15.3%) | 473 (15.0%) | 0.008 | 0.677 | 1394 (15.2%) | 375 (15.0%) | 0.008 | 0.734 |
| **EMS Prehospital Notification, n (%)** | 5617 (57.1%) | 2078 (57.3%) | 0.004 | 0.806 | 5981 (56.3%) | 1714 (60.2%) | 0.079 | <.001 |
| **Door-to-CT time, median [IQR]** | 19.0 [10 - 52] | 19 [10 - 46] | 0.014 | 0.482 | 19 [10 - 52] | 18.5 [10 - 45] | 0.006 | 0.789 |
| **Door-to-CT < 25 mins, n (%)** | 5720 (58.6%) | 2165 (60.5%) | 0.039 | 0.048 | 6173 (58.7%) | 1712 (60.8%) | 0.044 | 0.039 |
| **Discharge Home, n (%)** | 3549 (35.9%) | 1265 (34.5%) | 0.029 | 0.136 | 3803 (35.6%) | 1011 (35.0%) | 0.013 | 0.547 |

Note: SD = standard deviation; IQR = interquartile range; NIHSS = National Institutes of Health Stroke Scale; TIA = transient ischemic attack; NOS not otherwise specified; CT = computed tomography
Matched refers to cases successfully linked between EMS and hospital datasets, unmatched refers to cases appearing in hospital dataset not matched to corresponding EMS record

**Table 2.** Multivariable Logistic Re gression of Factors Associa ted with Successf ul Match: Determin istic vs. Probabilistic Approaches.

|  | Deterministic |  | Probabilistic |  |
| --- | --- | --- | --- | --- |
|  | aOR (95% CI) | p-value | aOR (95% CI) | p-value |
| **Age, mean (SD)** | 1.08 (1.03 - 1.14) | 0.001 | 1.08 (1.02 - 1.13) | 0.006 |
| **Sex, Female** | 0.97 (0.90 - 1.05) | 0.468 | 0.97 (0.89 - 1.06) | 0.535 |
| **Race, n (%) White** | 0.62 (0.54 - 0.70) | <.001 | 0.58 (0.51 - 0.67) | <.001 |
| **Insurance Status** |  | 0.029 |  | <.001 |
| Medicare | Reference |  | Reference |  |
| Non-Medicare | 1.08 (0.97 - 1.21) |  | 1.11 (0.98 - 1.25) |  |
| Missing | 1.19 (1.04 - 1.36) |  | 1.40 (1.21 - 1.63) |  |
| **Stroke type** |  | 0.051 |  | 0.046 |
| Ischemic Stroke/TIA | Reference |  | Reference |  |
| Hemorrhagic Stroke | 1.16 (1.03 - 1.32) |  | 1.18 (1.03 - 1.35) |  |
| Stroke NOS/No<br>Stroke Diagnosis | 0.95 (0.57 - 1.59) |  | 0.84 (0.49 - 1.44) |  |
| **Year** |  | <.001 |  | <.001 |
| 2021 | Reference |  | Reference |  |
| 2022 | 0.85 (0.77 - 0.93) |  | 0.87 (0.78 - 0.96) |  |
| 2023 | 0.77 (0.70 - 0.85) |  | 0.73 (0.65 - 0.80) |  |
| **NIHSS Category** |  | 0.003 |  | 0.007 |
| 0-5 | Reference |  | Reference |  |
| 6-10 | 0.94 (0.84 - 1.05) |  | 0.94 (0.84 - 1.06) |  |
| 11-15 | 0.84 (0.74 - 0.96) |  | 0.79 (0.68 - 0.91) |  |
| 16-20 | 0.86 (0.74 - 1.01) |  | 0.83 (0.70 - 0.99) |  |
| >20 | 0.84 (0.74 - 0.96) |  | 0.94 (0.81 - 1.09) |  |
| Missing | 0.75 (0.63 - 0.89) |  | 0.80 (0.67 - 0.96) |  |
| **EMS Prehospital**<br>**Notification** | 1.02 (0.94 - 1.11) | 0.622 | 0.88 (0.80 - 0.96) | 0.004 |
| **Door-to-CT Time** |  | 0.012 |  | 0.156 |
| ≤25 | Reference |  | Reference |  |
| >25 | 1.08 (0.99 - 1.18) |  | 1.04 (0.95 - 1.14) |  |
| Missing | 0.72 (0.53 - 0.97) |  | 0.76 (0.54 - 1.06) |  |
| **Discharge Home** | 1.03 (0.94 - 1.13) | 0.502 | 1.00 (0.91 - 1.10) | 0.941 |

Notes: aOR = adjusted odds ratio; CI = confidence interval; NIHSS = National Institutes of Health Stroke Scale; TIA = transient ischemic attack; NOS = not otherwise specified; CT = computed tomography
Age is scaled such that the odds ratio reflects a one standard deviation (14-year) increase in age.

Similar patterns emerged for the probabilistic matching strategy. There was a meaningful difference for race with a higher proportion of unmatched patients being White (90.6% vs. 85.2%; standardized difference = 0.164; p < 0.001). Admission year also differed (standardized difference = 0.072; p < 0.001), with unmatched patients more likely to be from 2023. NIHSS scores were higher among unmatched patients (median 6 vs. 5; standardized difference = 0.054; p = 0.014), and there was a modest difference in NIHSS category distribution (standardized difference = 0.034; p = 0.004,). Notably, EMS prehospital notification was more common in unmatched patients (60.2% vs. 56.3%; standardized difference = 0.079; p < 0.001). There were no differences for other variables, including age, sex, treatment, and outcomes (standardized differences < 0.05). It, therefore, does not appear that the higher match rate using the probabilistic method increased the representativeness of the sample.

The multivariable logistic regression model used to estimate selection probability is presented in Table 3. Effects were only interpreted when overall tests were p < .05. In these models, several characteristics were associated with the likelihood of a patient being matched, with results largely consistent between deterministic and probabilistic approaches.

**Table 3.** Adjusted Odds Ratios and Percent Change Comparing Determ inistic and Probabilistic Matching Methods Be fore and After Weighting for Selection Bias.

| Outcome | Matching Method | aOR (95% CI) | p-value | Weighted aOR (95% CI) | p-value | % Change in aOR |
| --- | --- | --- | --- | --- | --- | --- |
| **Door-to-CT** | Deterministic | 5.38 (4.85, 5.98) | <.001 | 5.35 (4.88, 5.85) | <.001 | −0.72% |
|  | Probabilistic | 5.11 (4.62, 5.65) | <.001 | 5.06 (4.62, 5.54) | <.001 | −0.58% |
| **Discharge Disposition** | Deterministic | 1.08 (0.98, 1.20) | 0.132 | 1.09 (1.00, 1.18) | 0.065 | 0.46% |
|  | Probabilistic | 1.09 (0.99, 1.20) | 0.069 | 1.10 (1.01, 1.20) | 0.033 | 0.46% |

Note: aOR = Adjusted odds ratio; CI = confidence interval
% Change in aOR is calculated as the relative change between the weighted and unweighted aOR

Older age was associated with increased odds of matching (Deterministic aOR = 1.08 (95% CI 1.03–1.14); Probabilistic aOR = 1.08 (95% CI 1.02–1.13), p = .001). White race was associated with lower odds of matching compared to non-White individuals in both models (Deterministic aOR =0.62 (95% CI 0.54-0.70); Probabilistic aOR = .058 (95% CI 0.51-0.67), p<0.001). Insurance status and year were also associated with matching in both models. Patients with missing insurance had higher odds of matching, whereas those from later years, particularly 2023, had lower odds relative to 2021. Stroke type was associated with matching in the probabilistic model (p = 0.046) but not in the deterministic model (p = 0.051). In the probabilistic model, hemorrhagic stroke was associated with higher odds of matching compared to ischemic stroke (aOR = 1.18 (95% CI 1.03-1.35), p = 0.018). NIHSS category was associated with matching in both models. Patients who had more severe strokes (NIHSS 11 or greater) or missing scores had a reduced odds of matching relative to those with mild strokes (NIHSS 0–5). EMS prehospital notification was associated with lower odds of matching only in the probabilistic model (aOR = 0.88 (95% CI 0.080-0.96), p = 0.004). Door-to-CT time was associated with matching in the deterministic (p = 0.012) but not in the probabilistic model (p = 0.156). Discharge destination was not associated with matching in either model.

### Potential Impact of Selection Bias

Weighted and unweighted results were similar for the impact of EMS identification of stroke on door-to-CT time. In the deterministic match model, the adjusted odds ratio decreased slightly from 5.38 to 5.35, a percent change of −0.72%. In the probabilistic match model, the adjusted odds ratio changed from 5.11 to 5.06, a percent change of −0.92% (each p < .001). Patients who were identified by EMS as having had a stroke were over 5 times more likely to have a door-to-CT time less than 25 minutes compared to those patients not identified as having a stroke by EMS.

The impact of adjusting for selection bias on the relationship between EMS identification and discharge home was similar. In the deterministic match model, the adjusted odds ratio increased from 1.080 to 1.085, a percent change of 0.46%. In the probabilistic match model, the adjusted odds ratio changed from 1.093 to 1.098, a percent change of 0.46%. This effect was not significant in the deterministic model (p = 0.065), but reached significance in the probabilistic model (p = 0.033), with patients who EMS identified as having a stroke being slightly more likely to be discharged home.

## Discussion

We compared deterministic and probabilistic matching methods for linking EMS and in-hospital stroke registry data in the absence of unique identifiers. Our main finding is that both approaches produced high match rates (73.0% for deterministic and 78.7% for probabilistic linkage). Matched samples from the two methods were similarly representative of the full GWTG-Stroke population transported by EMS, and differences in measured patient characteristics between matched and unmatched cases were modest across both linkage methods. Furthermore, the impact of accounting for selection bias introduced by incomplete matching on estimates of associations between EMS data and in-hospital treatment and outcomes was small.

Our match rates exceed those reported in similar linkage studies in stroke populations. For example, one study reported a 62.8% match rate using deterministic linkage in North Carolina stroke registry data.^18^ Another study conducted in Michigan compared these linkage methods and found match rates of 45.5% and 62.8% for deterministic and probabilistic approaches, respectively.^19^ Compared to these studies, the higher match rates in our study may reflect improved data quality, more consistent documentation, or better compatibility between EMS and hospital datasets. It may also reflect the more recent time frame of our study (2021– 2023), during which time electronic health and EMS record systems have become more standardized and interoperable. Our matching rates, however, are lower than those found in non-stroke settings. Several studies linking EMS records to in-hospital or hospital discharge records in the US and Canada found match rates ranging from 88% to 95% using deterministic or hierarchical deterministic algorithms.^22–24^ These studies were able to use personal identifying information such as names, social security numbers, and personal health identifiers, suggesting that the availability of these personal identifiers can lead to a meaningful increase in matching rates.

Although probabilistic linkage yielded a higher match rate, it did not meaningfully improve the representativeness of the linked sample, contrary to previous studies using similar data sources.^19^ In our data, matched patients under both methods were slightly younger, more likely to be from earlier years of the study period, and less likely to be White compared to unmatched patients. The differences by race and year persisted, even after probabilistic matching, highlighting that increasing match rates alone does not guarantee a more representative sample. Indeed, notable differences have been found between matched and unmatched records with matching rates as high as 95%.^24^ This is important, as probabilistic linkage is often assumed to be “better” because it includes more records; our findings suggest that this assumption should be critically evaluated in each setting.

When we examined the potential impact of selection bias on downstream analyses, we found that accounting for selection bias by using only the linked subset of the sample did not substantially alter the results. Odds ratio estimates were largely consistent before and after applying inverse probability weighting, suggesting that the impact of linkage-related selection bias on our analyses was minimal. These small differences can be attributed to the fact that both deterministic and probabilistic samples were representative of the overall sample, and that the probabilistic method did not greatly improve representativeness.

Together, these findings suggest that when linkage rates are high and differences between matched and unmatched samples are small, both deterministic and probabilistic methods yield valid and comparable results. In these situations, ease of implementing the deterministic match may override the increase in match rates afforded by the more time-consuming probabilistic method. Importantly, even in the absence of individual identifiers, useful and relatively unbiased analyses of EMS-hospital linked data are feasible, which supports ongoing efforts to use EMS registries and hospital data together to evaluate stroke systems of care.

Several limitations of our analysis should be noted. First, without access to a “gold standard” or unique identifiers, we could not validate our matches or directly quantify false positives and false negatives. As a result, we cannot assess the accuracy of either linkage method beyond indirect comparisons of representativeness and selection bias. Second, the study was conducted in a single state, potentially limiting generalizability. Patterns of EMS care, hospital documentation, and registry participation may differ in other regions, impacting the decisions to pursue deterministic vs probabilistic matching. Third, our study period overlapped with the COVID-19 pandemic, which may have affected EMS and hospital processes, patient characteristics, and outcomes in ways that could influence linkage or generalizability. Finally, although we adjusted for multiple confounders in the IPW models, it is likely that the variables used in the weighting model did not fully capture all relevant differences between matched and unmatched patients. If important unmeasured factors influenced both the likelihood of linkage and the outcomes, some residual bias may remain. As such, although our results suggest minimal impact from observed selection factors, the potential for residual selection bias due to unobserved characteristics remains and warrants further investigation in future studies. Lastly, hospital data was limited to facilities participating in the GWTG-S program. Similar findings may not be found in hospitals using different EMS or medical documentation systems.

With high match rates and similar representativeness, we found that both deterministic and probabilistic linkage methods produced comparable results in linking EMS and in-hospital stroke data. Although probabilistic methods modestly increased match rates, they did not substantially improve representativeness or reduce selection bias. Our findings suggest that, when high-quality data with low levels of missingness are available, deterministic linkage may be sufficient for many analyses, and the impact of linkage-related selection bias may be minimal. Even in these situations, however, it is important to perform sensitivity analyses to assess the potential impact of measured selection bias on the study’s findings when information on the full sample is available. As data systems become more integrated, continued efforts to facilitate secure and accurate linkage of EMS and hospital data will be essential for evaluating and improving stroke systems of care.

## Sources of Funding

This study was supported by the US Centers for Disease Control and Prevention as part of the Paul Coverdell National Acute Stroke Program (NU58DP006953). Get With The Guidelines-Stroke is funded by the American Heart Association and the American Stroke Association.

## Disclosures

This work represents the authors’ independent analysis of local or multicenter data gathered using the AHA Get With The Guidelines® (GWTG) IQVIA Registry Platform but is not an analysis of the national GWTG dataset and does not represent findings from the AHA GWTG National Program.

## Data Availability

Data for the current study are not publicly available due to data sharing agreements between participating hospitals and American Heart Association data use restrictions.

## Acknowledgements

The authors would like to thank Brent McKune and Kari Moore for their contributions to the conceptualization of the research questions addressed in this manuscript. Their efforts in securing funding that supported this work were instrumental in making this research possible.

## Notes

### Competing Interest Statement

The authors have declared no competing interest.

### Author Declarations

University of Kentucky Office of Research Integrity

